# Increased functional connectivity of thalamic subdivisions in patients with Parkinson’s disease

**DOI:** 10.1101/19002139

**Authors:** Conor Owens-Walton, David Jakabek, Brian D. Power, Mark Walterfang, Dennis Velakoulis, Danielle van Westen, Jeffrey C.L. Looi, Marnie Shaw, Oskar Hansson

## Abstract

Parkinson’s disease (PD) affects 2-3% of the population over the age of 65 with loss of dopaminergic neurons in the substantia nigra impacting the functioning of basal ganglia-thalamocortical circuits. The precise role played by the thalamus is unknown, despite its critical role in the functioning of the cerebral cortex, and the abnormal neuronal activity of the structure in PD. Our objective was to more clearly elucidate how functional connectivity and morphology of the thalamus are impacted in PD (*n* = 32) compared to Controls (*n* = 20). To investigate functional connectivity of the thalamus we subdivided the structure into two important regions-of-interest, the first with putative connections to the motor cortices and the second with putative connections to prefrontal cortices. We then investigated potential differences in the size and shape of the thalamus in PD, and how morphology and functional connectivity relate to clinical variables. Our data demonstrate that PD is associated with increases in functional connectivity between motor subdivisions of the thalamus and the supplementary motor area, and between prefrontal thalamic subdivisions and nuclei of the basal ganglia, anterior and dorsolateral prefrontal cortices, as well as the anterior and paracingulate gyri. These results suggest that PD is associated with increased functional connectivity of subdivisions of the thalamus which may be indicative alterations to basal ganglia-thalamocortical circuitry.

## Introduction

Parkinson’s disease (PD) is the second most common neurodegenerative disorder in the world, affecting 2-3% of the population over the age of 65 [1]. Characteristic motor symptoms of the disorder include resting tremor, rigidity and postural instability. These are accompanied by non-motor symptoms including cognitive impairment, autonomic dysfunction, disorders of sleep-wake cycle regulation, sensory disturbances and pain [2]. The neuropathological hallmark of PD is the presence of α-synuclein-immunopositive Lewy bodies and neurites [3]. This neuropathology results in a degeneration of nigrostriatal dopaminergic neurons, depletion of dopamine across the striatum [4] and consequent dysfunction of basal ganglia-thalamocortical networks [5, 6]. α-synuclein is thought to spread from brainstem regions along the neuraxis, impacting areas of the neocortex at advanced stages of the disease [7]. Dysfunction of basal ganglia-thalamocortical circuits is thus critical as these circuits work in concert with the cortex to mediate a range of cognitive, motor and limbic functions in the brain [8].

To understand the functioning of a network, it is necessary to study the elements of the network and also their interconnections [9]. Investigating elements of brain networks can be done via studying the morphology of key neuroanatomical nuclei acting as ‘hubs’ within these networks [10]. Hubs are nodes within brain networks that make strong contributions to global network function [11] and the interconnections between these hubs can be investigated through the use of resting-state functional connectivity MRI methods that measure the functioning of intrinsic connectivity networks in the brain at rest [12]. Spontaneous fluctuations of the blood oxygen level-dependent (BOLD) signal that are temporally coherent indicate areas in the brain that may be functionally and anatomically related [13].

As dopamine replacement therapies provide some relief of motor symptoms in PD, significant research effort has focused on the role played by dopaminergic-depleted nuclei of the striatum [14]. However, there is now considerable evidence that the pathology underlying PD affects certain nuclei in the thalamus, and that the structure plays an important role in PD as loss of dopaminergic input to the striatum results in increased GABA-mediated inhibition of thalamocortical projections [15]. The thalamus, considered an integrative hub within functional brain networks [16], is thus an important neuroanatomical structure which we can use to investigate how basal ganglia-thalamocortical circuits are affected in PD, potentially revealing important information about the pathophysiology of the disease.

Functional connectivity (FC) studies implicating the thalamus have yielded inconsistent results with research demonstrating both increased coupling between the thalamus and sensorimotor regions in PD [17] and no significant differences in thalamic FC in PD [18]. Studies have indicated that PD is associated with thalamic volumetric changes compared to control groups [19, 20], while other work presents conflicting data [21-27]. Research has demonstrated subtle morphological changes in a PD cohort compared to controls [19, 23] while other research has found no significant localized shape changes [21, 24, 26]. Although the thalamus plays a key modulatory role in the brain, there is a lack of evidence for a relationship between thalamus volumes and clinical function [21, 22, 27, 28]. Further, there is still significant information to be derived about the precise contribution of component nuclei of the structure, which are impacted differently by the pathological changes that take place in PD. We thus aimed to investigate the FC of relevant functional subdivisions within the thalamus that are crucial in mediating cognitive and motor function. We then sought to investigate possible localized structural changes to the thalamus using a morphological surface-based and volumetric analyses.

We hypothesized that there would be morphological alterations to the thalamus in PD based on the potential impact of PD neuropathology, which has been shown to display neurodegeneration post-mortem [29]. We also hypothesized that there would be a correlation between smaller overall volumes of the thalamus and poorer performance on measures of motor and cognitive function. Finally, we hypothesized that FC of important motor and prefrontal subdivisions of the thalamus would be impacted in PD due to the effect of PD disease processes on basal ganglia-thalamocortical circuits.

## Materials and methods

### Participants

Participants in this research were members of the Swedish BioFINDER Study (www.biofinder.se). The study is based in Sweden and is affiliated with the Clinical Memory Research Unit and The Biomedical Centre, both at Lund University. Participants were recruited from the Memory and neurology clinics at Skåne University Hospital as well as the Memory Clinic at Ängelholm Hospital. Participants gave written informed consent and this research was performed in accordance with the World Medical Association’s Declaration of Helsinki. Participants in the current study received their clinical assessments between 23/05/2012 and 13/03/2014. Ethical approval was obtained through the Ethical Review Board of Lund, Sweden, and the Human Research Ethics Committee at the Australian National University, Canberra, Australia. Diagnosis of PD (*n* = 32) was made by a neurologist using the National Institute of Neurological and Stroke Diagnostic criteria [30]. A healthy control group (Controls) (*n* = 20) was used for comparison. Exclusion criteria for the Swedish BioFINDER study included poor knowledge of the Swedish language, developmental disability, psychiatric disorder, alcohol or substance abuse, the presence of a metabolic disorder, diagnosis of probable PD dementia [31]. A preliminary investigation of functional MRI data indicated a significant difference in subject head motion during image acquisition. We therefore implemented a strict study-specific head motion exclusion criterion of > 0.26mm (defined as mean relative displacement) and used advanced denoising procedures (FSL-FIX) as nuisance regression can be insufficient in removing the spurious effects of movement artifacts in MRI data [32].

All participants underwent a cognitive and neurological examination by a medical doctor with extensive experience in movement disorders. PD patients remained on medication as per their usual regime for both MRI acquisition and clinical assessment, with a levodopa equivalent dosage (LED) metric recorded for each participant. Functioning of participants was quantified with the Unified Parkinson’s Disease Rating Scale Part-III test (UPDRS-III), assessing the motor signs of PD [33], the Mini Mental State Examination (MMSE), assessing cognitive mental state [34], the Timed Up and Go test (TUG), assessing mobility [35], the A Quick Test of Cognitive Speed test assessing perception and cognitive speed (AQT) [36] and the Animal Fluency test (AF), assessing verbal fluency and executive function [37].

### MRI acquisition

Magnetic resonance imaging was performed on a 3T scanner (Trio, Siemens Magnetom, Erlangen, Germany) equipped with a 20-channel head-coil. High-resolution T1-weighted three-dimensional anatomical brain images were acquired using a magnetization-prepared rapid acquisition technique with gradient-echo sequence (repetition time = 7 ms; echo time = 3 ms; flip angle = 90 degrees; voxel size = isotropic 1=mm^3^). Image matrix size was 356 voxels in the coronal and sagittal planes and 176 voxels in the axial plane. Resting state functional magnetic resonance images (rs-fMRI) (256 volumes per subject) were acquired using T2*-weighted echo planar imaging volumes (repetition time = 1850 ms; echo time 30 ms; flip angle = 90 degrees; matrix 64 × 64; voxel size 3 × 3 × 3.75 mm^3^). Image matrix size was 64 voxels in the coronal and sagittal planes and 36 voxels in the axial plane. Subjects were instructed to lie still with their eyes closed, not to fall asleep and not to think of anything in particular during the scan, which lasted for approximately 8 minutes.

### Manual segmentation of the thalamus

Manual region-of-interest tracing was performed on participant’s T1 MRI data, using ANALYZE 12.0 software (Mayo Biomedical Imaging Resource, Rochester, Minnesota, USA) following a validated method [38]. The tracing for each thalamus was saved as a binary image for rs-fMRI seed-based FC and shape-based morphological analyses. A detailed explanation of the manual tracing method and associated reliability statistics is available in Supplementary Information ‘*Manual segmentation of the thalamus*.’

### Resting-state fMRI preprocessing

All rs-fMRI preprocessing used FMRIB Software Library (FSL) software package tools (FMRIB Software Library, Oxford, UK; FSL version 5.0.10, RRID:SCR_002823) [39]. FMRI Expert Analysis Tool (FEAT) (version 6.00) was used for the removal of the first 6 volumes, motion correction using FMRIB Motion Correction Linear Registration Tool (MCFLIRT) [40], slice-timing correction using Fourier-space timeseries phase-shifting, removal of non-brain structures using FSL’s Brain Extraction Tool (BET) [41], spatial smoothing using a full-width half-maximum gaussian kernel of 5 mm, grand mean intensity normalization and high-pass temporal filtering (gaussian-weighted least-squares straight line fitting, with sigma = 50.0s) [42]. Registration of functional images to subjects structural images used boundary-based registration [43] within the FMRIB Linear Registration Tool (FLIRT) [40, 44]. Registration of functional images to Montreal Neurological Institute (MNI) 152 T1 2mm^3^ standard space was also performed using FLIRT with 12 degrees of freedom, further refined using FNIRT nonlinear registration [45, 46] with a warp resolution of 10mm and a resampling resolution of 4mm. Denoising of head motion, scanner and cerebrospinal fluid artefacts was performed using a probabilistic Multivariate Exploratory Linear Optimized Decomposition into Independent Components (MELODIC version 3.15) independent component analysis method [47]. FSL’s ICA-based Xnoiseifier (FIX version 1.06) [48] was then used to classify components as either signal or noise, with noise components and also motion confounds (24 regressors) regressed from the data. For the full explanation of the denoising procedure see Supplementary Information ‘*Independent component analysis denoising*.’

The Oxford Thalamic Structural Connectivity Probability Atlas [49] within FSL’s visualization GUI FSLeyes was used to parcellate bilateral thalamic manual segmentation masks into two seed regions-of-interest masks (seed-ROIs) (Fig 1), representing important functional subdivisions of the thalamus. Due to the cardinal motor symptoms in PD, our first seed-ROI incorporated voxels with the highest probability of connectivity with pre- and primary motor cortices and are intended to represent the ventral lateral posterior, ventral lateral and ventral anterior thalamic nuclei. Due to the significant cognitive dysfunction observed in PD, our second seed-ROI incorporated voxels with the highest probability of connectivity to the prefrontal cortex, intended to represent the mediodorsal and anterior thalamic nuclei [49]. For ease of reference we will refer to these seed-ROIs as the VLp/VA thalamus and MD/A thalamus, respectively. Generic VLp/VA and MD/A thalamic masks were thresholded to only include voxels that had a greater than 50% chance of inclusion, then registered to subject-specific functional MRI space, and finally eroded by zeroing non-zero voxels when found in kernel, to reduce partial volume effects. A visual inspection of a subset of VLp/VA and MD/A masks was then performed to check the alignment of masks within functional data. We then extracted the mean activation of the functional data within the two seed-ROI masks at each functional timepoint for use as explanatory variables at the individual-level general linear model (GLM) stage in a mass univariate voxelwise whole-brain analysis. The average number of voxels per seed-ROI for each group and a pairwise comparison of this data is shown in Supplementary Information ‘*Atlas-based ROI segmentation statistics*.’

**Fig 1.**
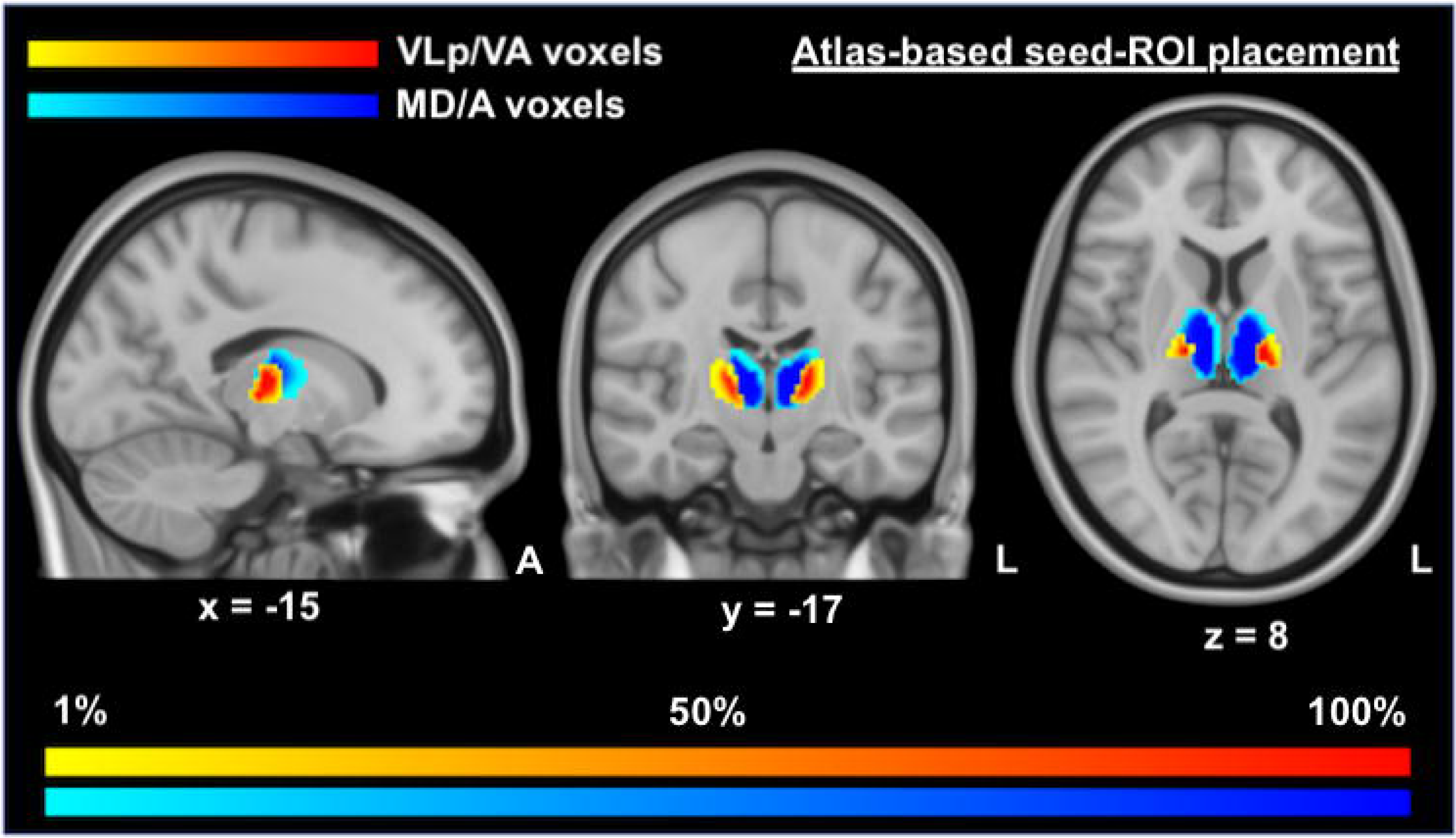
Positioning and likelihood-map of seed voxels for VLp/VA and MD/A thalamic masks. This figure displays the voxels used in seed-ROI masks for the functional connectivity analyses, overlaid on MNI T1 0.5mm images. This image was produced by combining all of the binary masks of each participant into one unified mask, with warm (yellow - red) colors indicating the positioning of the VLp/VA voxels and cold colors (light blue - dark blue) indicating the positioning of the MD/A voxels. Darker tones are indicative of a greater proportion of voxels in that region being included in the relevant seed-ROI mask. VLp/VA, ventral lateral posterior and ventral anterior thalamic voxels; MD/A, mediodorsal and anterior thalamic voxels.

### Resting-state fMRI statistical analyses

Two individual-level FC analyses were performed for each participant, using a GLM approach [50]. BOLD timeseries data within the VLp/VA and MD/A thalamus were correlated with activity in the rest of the brain, shifting the model with a temporal derivative to account for lags in the onset of the hemodynamic response function, removing volumes corrupted by large motion and regressing out average timeseries data from whole brain (global signal regression), white matter and ventricle masks. For the full explanation of our approach to individual-level GLM analyses, see Supplementary Information ‘*Individual-level GLM functional connectivity analyses*.’ Higher-level analysis of FC differences between PD and Control subjects were investigated in standard space using FSL-FEAT. We chose a nonparametric permutation-based approach (n = 5000) [51] via FSL-randomise [52] with a threshold-free cluster enhancement method controlling the family-wise error rate at *p* < 0.05. This approach avoids selecting arbitrary threshold values, while also potentially improving sensitivity to test signal shapes and SNR values [53]. Due to there being a significant difference in education between PD and Control groups, we included years of education as a covariate, along with age and sex.

### Correlation between functional connectivity and clinical data

To investigate the relationship between FC and clinical variables we conducted a series of post-hoc partial correlational analyses. These focused on the average parameter estimate for VLp/VA and MD/A thalamus at the peak voxel locations derived from the between group analyses. Spherical ROIs (7mm radius) were generated around the voxel locations with the average parameter estimate extracted for each subject. Average FC for each ROI was then correlated against LED, disease duration, UPDRS-III, TUG, AQT and Animal Fluency scores.

### Morphology of the thalamus in PD

#### Volumetric analyses

To investigate differences in volumes of the thalamus between PD and Controls we used SPSS 22.0 (IBM Corporation, Somers, New York, USA) utilizing multivariate analysis of covariance models controlling for estimated total intracranial volume (eTIV, derived from recon-all FreeSurfer processing [54]), age and sex. Effect sizes are represented by partial eta squared values (η^2^).

#### Surface based shape analysis

Shape analysis was performed using spherical harmonic parameterization and sampling in a three-dimensional point distribution model (SPHARM-PDM) [55] outlined in detail in Supplementary Information ‘*SPHARM-PDM detail*.’ Generally speaking, SPHARM-PDM shape analysis provides visualizations of the local surface changes to the thalamus between groups via mean difference displacement maps, mapping the magnitude of surface change (deflation or inflation) in millimeters between corresponding points on the mean surfaces of the PD subjects compared to Controls. Significant surface change was displayed at p < 0.05 with a correction for multiple comparisons performed using a false-discovery rate bound *q* of 5% [56].

### Correlations between thalamic volumes and clinical symptoms

We used hierarchical multiple regression models to assess whether thalamic volumes can predict clinical symptoms. These models incorporated two levels, the first level controlled for eTIV, age, sex and years of education (the latter when dealing with measures of cognitive function), and the second level held the independent variable of interest (volume of right or left thalamus), measuring the unique contribution of that variable in predicting each measure of clinical function. Effect sizes are represented by standardized beta values (β).

## Results

### Participant characteristics

There were no significant differences in age, head size (eTIV) or proportions of males and females in groups, between the PD cohort and Controls. Years of education, UPDRS-III and AQT performance was significantly reduced in PD compared to Controls (Table 1).

**Table 1.** Demographic and clinical characteristics of participants.

| Item | Controls | PD | *p*-value |
| --- | --- | --- | --- |
| Number of participants | 20 | 32 | - |
| Female/Male | 10/10 | 18/14 | 0.878 |
| Age | 69.06 ± 6.86 | 69.36 ± 5.82 | 0.868 |
| LED | - | 523.36 ± 295.31 | - |
| Disease duration | - | 5.16 ± 3.62 | - |
| eTIV (cm<sup>3</sup>) | 156.81 ± 15.84 | 152.06 ± 17.71 | 0.333 |
| Years of education | 13.14 ± 3.68 | 9.78 ± 6.05 | 0.042 |
| Relative displacement | 0.13 ± 0.06 | 0.16 ± 0.06 | 0.066 |
| H&Y | - | 1.75 ± 0.55 | - |
| UPDRS-III | 2.40 ± 3.02 | 10.69 ± 7.06 | <0.001 |
| TUG | 8.60 ± 1.54 | 9.68 ± 2.39 | 0.079 |
| MMSE | 28.8 ± 1.2 | 28.28 ± 1.42 | 0.18 |
| AQT | 57.25 ± 15.09 | 66.47 ± 14.78 | 0.035 |
| AF | 24.80 ± 6.59 | 23.03 ± 5.67 | 0.309 |

Data presented as mean ± standard deviation; *p*-values, one-way independent samples *t*-test, excluding male/female numbers which was analyzed via a chi-square test for independence; LED, levodopa equivalent dosage (mg); eTIV, estimated total intracranial volume; Relative displacement, mean value derived from MCFLIRT FSL motion correction; H&Y, Hoehn and Yahr Scale; UPDRS-III, Unified Parkinson's Disease Rating Scale part III; TUG, Timed Up and Go test; MMSE, Mini Mental-state Examination; AQT, A quick test of cognitive speed; AF, Animal fluency test.

### Functional connectivity of the VLp/VA thalamus in PD

Analysis of the VLp/VA thalamus in PD found significant clusters of increased FC with the right supplementary motor area (BA6) and the left paracingulate gyrus (BA32). Analysis of the VLp/VA thalamus also found a significant cluster of decreased FC with the left lateral occipital cortex (BA19) (Fig 2, Table 2).

**Table 2.** Regions showing functional connectivity differences with the VLp/VA thalamus in PD.

| FC Cluster | Peak | Brain regions | Voxels | MNI |  |  |
| --- | --- | --- | --- | --- | --- | --- |
|  |  |  |  | x | y | z |
| Increased | 1 | R Supplementary motor area (BA6) | 64 | 12 | -2 | 42 |
|  | 2 | L Paracingulate gyrus (BA32) | 11 | -4 | 10 | 42 |
|  | 3 | R Supplementary motor area (BA6) | 10 | 10 | 6 | 44 |
| Decreased | 4 | L Lateral occipital cortex (BA19) | 18 | -30 | -86 | 24 |
|  | 5 | L Lateral occipital cortex (BA19) | 10 | -26 | -66 | 46 |
|  | 6 | L Lateral occipital cortex (BA19) | 3 | -26 | -68 | 56 |

Brain regions and associated coordinates represent significant peaks within each cluster ( $p < 0.05$ ). Abbreviations: FC, functional connectivity; L, left hemisphere; R, right hemisphere; BA, Brodmann area; MNI, coordinates for location of peak voxels in Montreal Neurological Institute 152 T1 2mm space. Labelling of brain regions based on the Harvard-Oxford Cortical/Subcortical Atlases.

**Fig 2.**
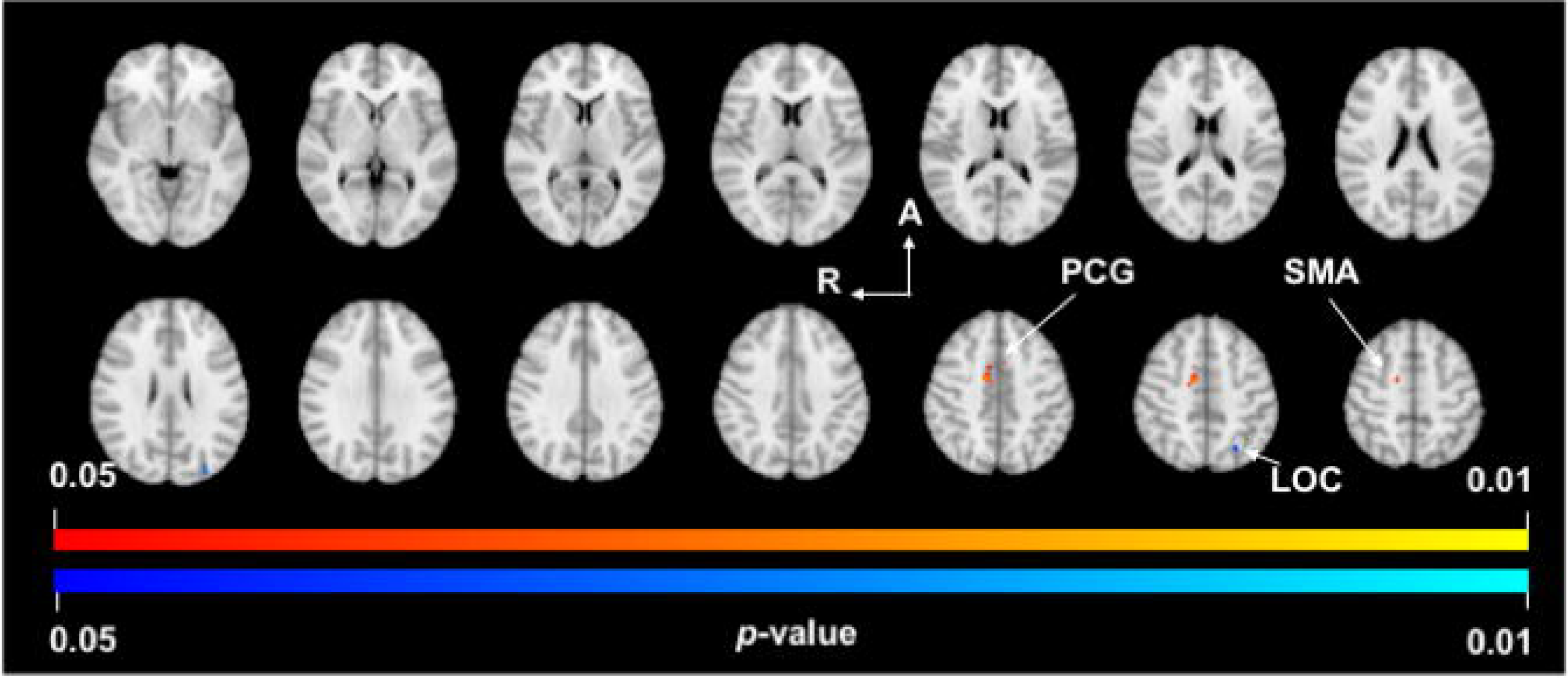
VLp/VA thalamus functional connectivity. *p*-value images showing neuroanatomical regions with significant between-group changes in functional connectivity with the VLp/VA thalamus in PD subjects compared to Controls. Warm colors (yellow-orange) represent areas of increased functional connectivity in PD and cool colors (light-dark blue) represent areas of decreased functional connectivity in PD. Spacing between each slice in the z-direction is 4.2mm beginning at z = −3.18 in the top left slice (MNI T1 2mm image). R, right; A, anterior; PCG, paracingulate gyrus; SMA, supplementary motor area; LOC, lateral occipital cortex.

### Functional connectivity of the MD/A thalamus in PD

Analysis of the MD/A thalamus in PD found significant clusters of increased FC with the left anterior cingulate (BA24) and left putamen (Fig 3, Table 3). These clusters extended across the following brain regions: the bilateral anterior (BA24) and paracingulate gyri (BA32), left caudate nucleus, left putamen, left globus pallidus, bilateral dorsolateral prefrontal cortex (BA9) and bilateral anterior prefrontal cortex (BA8). Analysis of the VLp/VA thalamus also found a significant cluster of decreased FC with the left lateral occipital cortex (BA19).

**Table 3.** Regions showing functional connectivity differences with the MD/A thalamus in PD.

| FC Cluster | Peak | Brain regions | Voxels | MNI |  |  |
| --- | --- | --- | --- | --- | --- | --- |
|  |  |  |  | x | y | z |
| Increased | 7 | L anterior cingulate (BA24) | 4319 | -8 | 18 | 22 |
|  | 8 | L putamen (BA49) | 268 | -20 | 10 | -4 |
| Decreased | 9 | L lateral occipital cortex (BA19) | 10 | -18 | -78 | 52 |

Brain regions and associated coordinates represent significant peaks within each cluster ( $p < 0.05$ ). Abbreviations: FC, functional connectivity; L, left hemisphere; R, right hemisphere; BA, Brodmann area; MNI, coordinates for location of peak voxels in Montreal Neurological Institute 152 T1 2mm space; Labelling of brain regions are based on the Harvard-Oxford Cortical/Subcortical Atlases.

**Fig 3.**
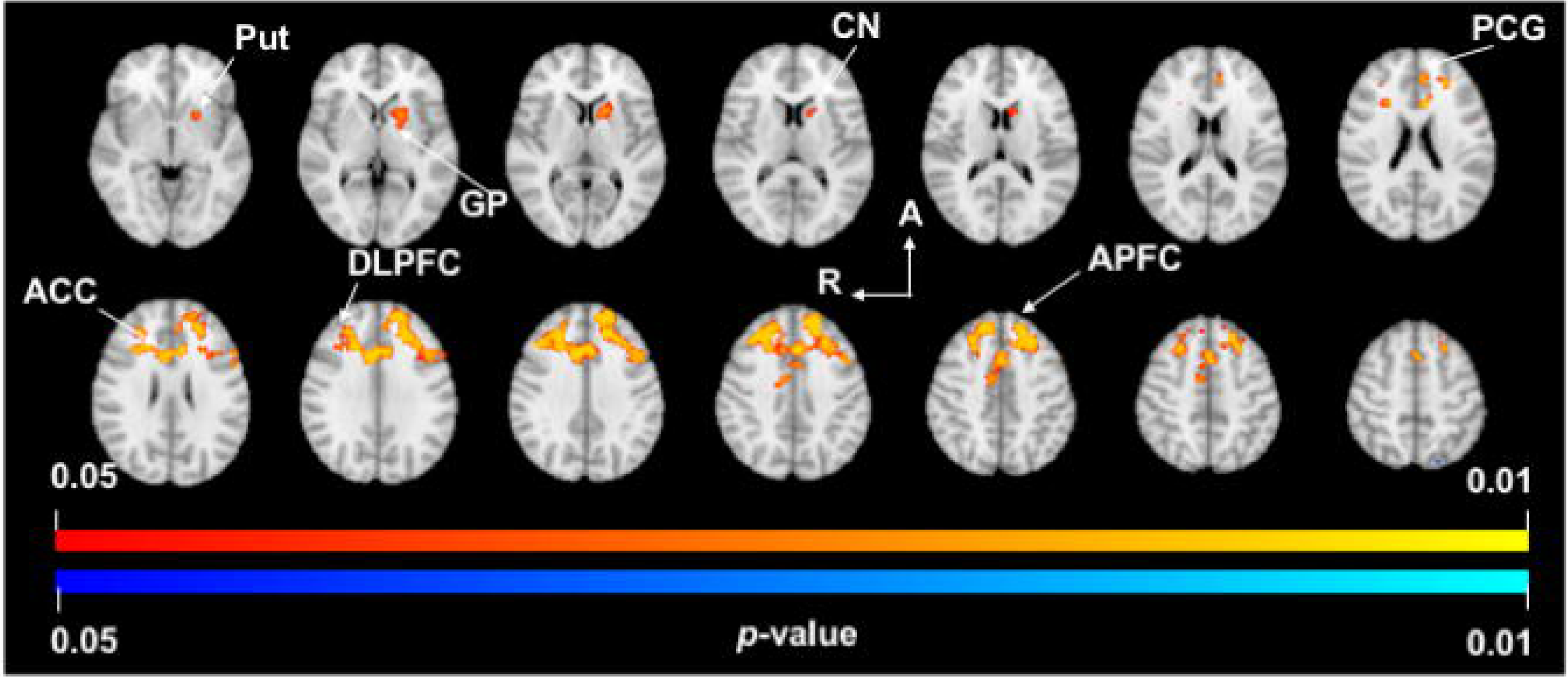
MD/A thalamus functional connectivity. *p*-value images showing neuroanatomical regions with significant between-group changes in functional connectivity of the MD/A thalamus in PD subjects compared to Controls. Warm colors (yellow-orange) represent areas of increased functional connectivity in PD. Spacing between each slice in the z-direction is 4mm beginning at z = −2.3 in the top left slice (MNI T1 2mm image). R, right; A, anterior; Put, putamen; GP, globus pallidus; CN, caudate nucleus; PCG, paracingulate gyrus; ACC, anterior cingulate cortex; DLPFC, dorsolateral prefrontal cortex; APFC, anterior prefrontal cortex.

### Correlation between functional connectivity and clinical data

In PD patients we observed a positive correlation between LED and mean FC of the right supplementary motor area (peak 3; r = 0.46, *p* = 0.01) and a negative correlation with the left lateral occipital cortex (peak 4; r = −0.41, *p* = 0.04). We also observed a positive correlation between disease duration and mean FC of the right supplementary motor area (peak 1; r = 0.41, *p* = 0.03; peak 3; r = 0.57, *p* = 0.01). We also observed a positive correlation between TUG scores and mean FC of the left paracingulate gyrus (peak 2; r = 0.43, *p* = 0.02) and lateral occipital cortex (peak 6; r = 0.43, *p* = 0.02; peak 9; r = 0.37, *p* = 0.047). None of these results survived correction for multiple comparisons (*Supplementary Information Table S1*).

### Morphology of the thalamus in PD

#### Volumetric analyses

Comparisons of thalamic volumes found no difference between the PD group and Controls (Table 4).

**Table 4:** Estimated mean volumes of right and left thalamus and pairwise comparison.

| Structure | Control | PD | Mean difference | *p*-value |
| --- | --- | --- | --- | --- |
| Right thalamus volume (mm<sup>3</sup>) | 5774.13 | 5927.45 | -153.32 | 0.25 |
| Left thalamus volume (mm<sup>3</sup>) | 5594.47 | 5749.24 | -154.78 | 0.18 |
| Estimated volumes of the right and left thalamus after adjusting for age, eTIV and sex. *p*-value presented has been adjusted for family-wise error rate using a Bonferroni correction. |  |  |  |  |

#### Surface based shape analysis

Shape analysis found no localized areas of shape change to the surface of the right or left thalamus in the PD group compared to Controls, after correcting for false-discovery rate (Fig 4).

**Fig 4.**
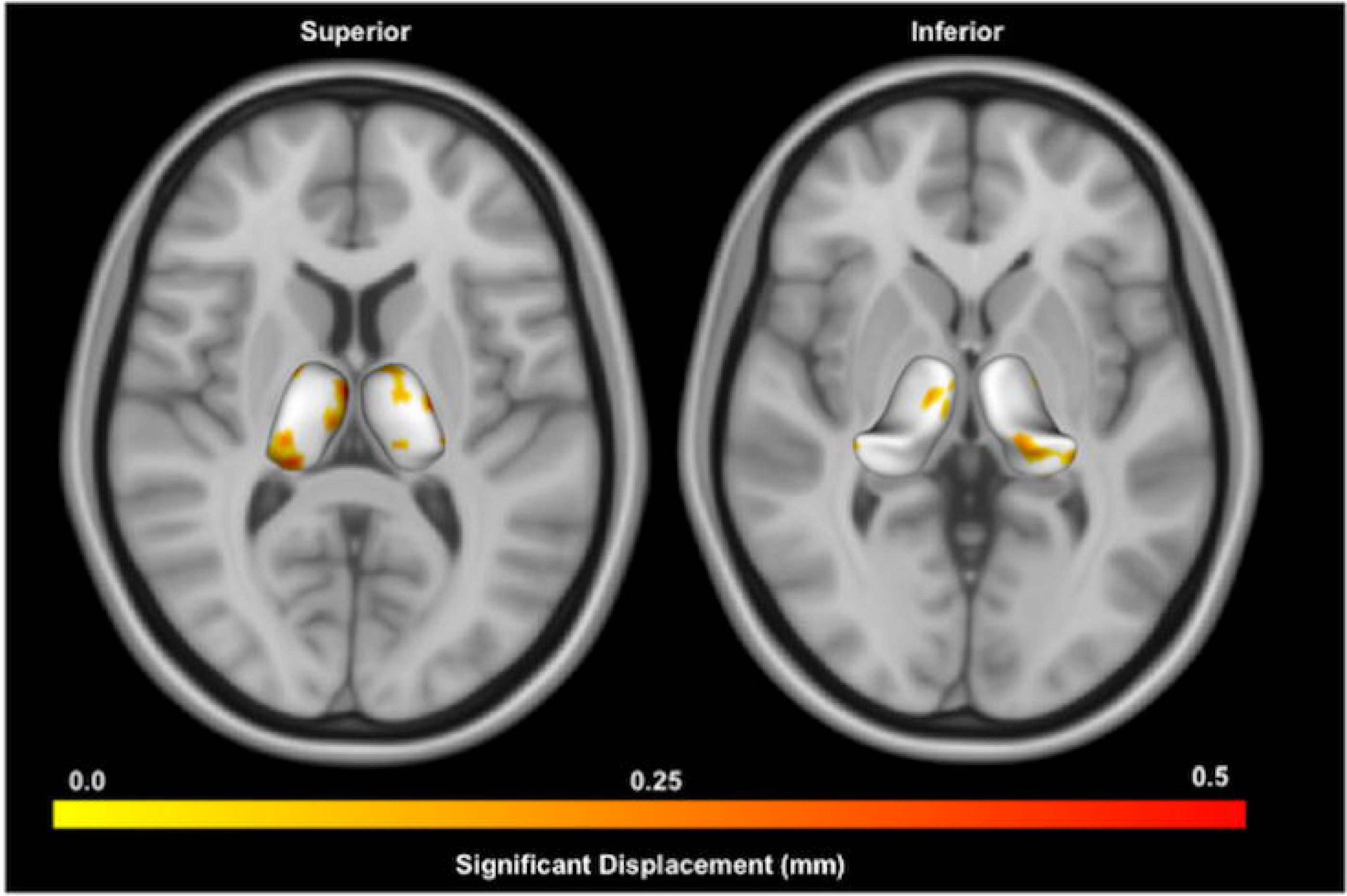
Shape analysis of thalamus in PD compared to Controls. Displayed are superior and inferior views of bilateral thalami overlaid on axial MNI T1 0.5mm images. Warmer colors indicate regions of greater inflation in the PD group compared to Controls using point-wise significance tests (*p* < 0.05, uncorrected). No regions were significant after false-discovery rate correction.

### Correlations between thalamic volumes and clinical symptoms in PD

We found no significant relationships between volumes of the right or left thalamus and clinical function in the PD or Control groups (*Supplementary Information Table S2*).

## Discussion

The results of this study demonstrate that PD patients on medication have increased FC within motor, dorsolateral and anterior cingulate basal ganglia-thalamocortical circuits. Our data support the findings of a recent meta-analysis showing that PD patients have increased FC of basal ganglia-thalamocortical circuity [57], and extend these findings by showing how important functional subterritories of the thalamus are impacted in PD. These changes in FC were found despite any evidence of morphological alterations to the thalamus.

Our findings of increased FC between the VLp/VA thalamus and the supplementary motor area may be indicative of changes within one segment of the classic ‘motor’ basal ganglia-thalamocortical circuit [5]. In this model, the motor circuit originates at the supplementary motor area, receiving input from the primary and the premotor cortices. These areas connect with the nuclei of the basal ganglia, which project back to the ventral anterior and ventral lateral nuclei of the thalamus, closing the loop by reconnecting with the site of origin in motor cortical areas [5]. Increased FC between the putamen and the supplementary motor area has been demonstrated previously [58] while other research using graph theoretical analyses has demonstrated increased functional connectivity within the sensorimotor network in PD [59]. Evidence suggests that increased FC of sensorimotor networks is likely related to dopaminergic medication usage [60] potentially facilitating increases in FC, which is a mechanism that will be discussed momentarily.

Our research also demonstrates significant and far more widespread increases in FC in PD between the MD/A thalamus and the anterior and dorsolateral prefrontal cortices, potentially indicating a disease related change to the ‘dorsolateral prefrontal’ basal ganglia-thalamocortical circuit [5]. This circuit originates in Brodmann areas 9 and 10 on the lateral surface of the anterior frontal lobe, and after traversing the basal ganglia, connects directly with our intended seed-ROIs at the anterior and mediodorsal nuclei of the thalamus. From there, the circuit projects back to the anterior and dorsolateral prefrontal cortices to form a closed loop [5]. Our findings of increased FC between the MD/A thalamus and the dorsolateral prefrontal cortex may represent functional compensatory mechanisms due to the significant cognitive dysfunction common in PD. The dorsolateral prefrontal cortex helps to execute tasks that contribute to cognitive functioning, including working memory, decision-making and action control, achieved through a top-down modulation of behavior in concert with diverse cortical and subcortical structures [61]. Research has shown that FC is significantly increased *across* the prefrontal cortex in PD subjects on medication, as the brain potentially recruits new anatomical areas to aid in the performance of cognitive tasks. It is argued that changes in FC may indicate a functional compensation to help restore cognitive processes in PD [61]. Interestingly, we also found significant increases in FC between the MD/A thalamus and the anterior cingulate cortex in PD subjects, indicating a disease related change to the ‘anterior cingulate’ basal ganglia-thalamocortical circuit. In this circuit, the anterior cingulate cortex links with the ventral basal ganglia structures, outputs to the ventral anterior nuclei of the thalamus, and links back with the anterior cingulate cortex [5]. Our findings of increased FC with the anterior as well as paracingulate gyri fit with the concept of increased FC due to compensatory mechanisms, as this region of the brain interacts with the lateral prefrontal cortex to mediate performance in tasks linked to cognitive processes [62]. Research has shown that a common response to neurological disruption is the *hyper*-connectivity of brain circuits, which may reflect a protective mechanism in the brain to maintain normal functioning [63]. Such a mechanism has been proposed in PD previously [64, 65], as well as in mild cognitive impairment and Alzheimer disease [66, 67], and taken together, our results provide support for this model.

There are nonetheless inconsistencies with similar FC research that require consideration. Our data contrast with work demonstrating no significant FC changes between the thalamus and widescale brain networks in subjects on medication [18]. A possible explanation for this inconsistency may relate to how FC changes across different disease stages. When compared to the current work, the study in question focused on PD subjects with both a longer disease duration as well as a higher average Hoehn and Yahr stage [18]. This is important because research has shown that FC in PD may undergo periods of both *hyper*-connectivity and *hypo*-connectivity as the disease progresses [64]. Potential FC changes across the course of PD thus make difficult to compare studies with subjects at different disease stages.

Our data also indicate that PD is associated with increases in FC between the MD/A thalamus and the left dorsal caudate nuclei, anterior putamen and globus pallidus. Strong evidence suggests that the output of the basal ganglia, mainly the globus pallidus interna, is hyperactive in PD [68] and our results of increased FC with this area suggest that increased activity may be accompanied by increased FC. Increased FC between the caudate nuclei and the thalamus has been shown in a PD cohort on medication [69], however two studies have demonstrated conflicting results [17, 70]. A crucial difference separating these studies relates to patient inclusion. One of these studies focused on early PD patients with a mean disease duration of 1.7 years [17], which differs markedly from the current study where the average disease duration is 5.16 years. The present study also excluded PD subjects with dementia, fundamentally distinguishing it from our previous work [70]. This is crucial as PD dementia is associated with decreases in FC compared to subjects without the diagnosis [71]. Our findings of increased FC between the MD/A thalamus and the anterior putamen are supported by similar research in this field [65]. This research demonstrated that increased inter-regional coupling of the anterior putamen, the region anterior to the anterior commissure, follow the specific spatial pattern of dopamine depletion in PD [4]. This research suggests that PD patients may undergo a shift in cortico-striatal connections from the neuro-chemically more affected posterior putamen toward the relatively spared anterior putamen. Our results support this finding, suggesting that the anterior putamen may undergo increased FC with the MD/A thalamus, further supporting the model that the pathophysiology of PD may involve compensatory alterations in the FC of key nuclei within basal ganglia-thalamocortical circuits.

While conceiving of FC changes in terms of segregated basal ganglia-thalamocortical circuits is attractive, this inference is theoretical. It is therefore helpful to consider the results of other imaging techniques to substantiate our findings. Graph-based eigenvector centrality mapping research (which informs on the number and quality of node connections within networks) has shown that this metric is increased in the thalamus in PD [72]. Interestingly, we also found a significant (though small) cluster of reduced FC of both the VLp/VA and MD/A thalamus with the lateral occipital cortex in PD patients, supporting previous work which found reductions in FC between nuclei of the extended basal-ganglia (including the thalamus) and this area of the brain [72]. The findings from ^18^F-flurodeoxyglucose PET imaging has indicated that PD is associated with increased pallidothalamic activity [73], which supports our findings. However, this research also demonstrated that regions of the dorsolateral prefrontal cortex and the supplementary and premotor areas show reductions in metabolic activity in PD, contrasting with our results [73].

We found no significant correlation between FC of our seed-ROI in the thalamus and measures of clinical function, disease duration or antiparkinsonian medication use. This supports previous meta-analytic research which indicates that FC within a basal-ganglia networks does not correlate with clinical indices of disease severity [74], arguing instead that altered FC reflects a constitutional alteration of the networks under consideration. Our results are also bolstered by meta-analytic findings which indicate that increased FC of the thalamus were unaffected by medication status [57].

The morphological data from our study are consistent with previous reports indicating that PD is not associated with atrophy of the thalamus [21, 23-28]. We also found no significant localized surface shape changes in the PD group, supporting a number of studies [21, 24, 25]. After investigating between-group morphological differences we investigated potential relationships between volumes of the thalamus and measures of clinical function. These analyses revealed no significant findings, supporting previous research [21, 27, 28]. These results make intuitive sense, as we found no significant volumetric or localized shape changes in the PD cohort, suggesting there was no discernable relationship between atrophy of the thalamus and the PD disease process.

### Strengths and Limitations

Possible limitations of the current work warrant further attention. The first is our small sample size. While this is an important factor that negatively impacts the power of our study, our sample size was the result of choosing a highly stringent head motion exclusion criteria, which we believe is a strength of our work. A second possible limitation is the use of atlas-based seed-ROI segmentation, defined by the structural connectivity of thalamic nuclei [49]. Future work may benefit from a data-driven parcellation scheme as it may better capture functional boundaries of seed-ROIs. Despite these considerations we believe our data make important statements about the role of the thalamus within basal ganglia-thalamocortical circuits in PD.

## Conclusions

We found increases in functional connectivity between the VLp/VA thalamus and the supplementary motor area and paracingulate gyrus, and also between the MD/A thalamus and basal ganglia nuclei, anterior and paracingulate cingulate gyri, anterior and also dorsolateral prefrontal cortical regions. Significant increases in functional connectivity were found despite any observable volumetric or localized shape alterations to the thalamus. The results of this study indicate that functional connectivity changes occur in PD, which likely result from disease-related system level dysfunction of the thalamus as a crucial network hub within basal ganglia-thalamocortical circuits.

## Data Availability

Anonymized data will be shared by request from any qualified investigator for the sole purpose of replicating procedures and results presented in the article and as long as data transfer is in agreement with EU legislation on the general data protection regulation. Data underlying the results described in our manuscript are available from at Attention: Conor Owens-Walton Academic Unit of Psychiatry & Addiction Medicine Australian National University Medical School Building 4, Level 2, Canberra Hospital Woden, A.C.T. 2605 AUSTRALIA or

## Acknowledgements

The authors are indebted to all patients and control subjects who participated in this study. This project is an initiative of the Swedish BioFINDER Study, of whom DvW and OH are steering committee members, and also the AUSSIE network coordinated by JCLL at the Australian National University Medical School, who self-funds related expenses.

## Supporting Information captions

**Supplementary Table S1. Correlations between functional connectivity and clinical data**. All values are rounded to two decimal places. VLp/VA thalamus, clusters 1-6, MD/A thalamus clusters 7-9. Significance based on a two-tailed Pearson correlation controlling for age, sex and years of education. A correction for multiple comparisons using the Bonferroni method stipulate a *p*-value of < 0.000925 required for significance (based on performing 54 analyses). LED, levodopa equivalent dosage; UPDRS-III, Unified Parkinson’s Disease Rating Scale part III; TUG, Timed Up and Go test; AQT, A quick test of cognitive speed test; AF, Animal fluency test.

**Supplementary Table S2. Correlations between thalami volumes and clinical measures: PD and Controls**. A correction for multiple comparisons using the Bonferroni method stipulate a *p*-value of < 0.00625 required for significance (based on performing 8 analyses). UPDRS-III, Unified Parkinson’s Disease Rating Scale part III; TUG, Timed up and Go test; AQT, A Quick Test of Cognitive Speed; AF, Animal Fluency test; R^2^ change, variance in clinical measure score explained by unique contribution of the volume of interest (multiply by 100 to find percentage); β, standardized beta coefficient, indicating effect size.

